# Nature contact by window opening and depressive symptoms among Older Adults: A cross-sectional analysis of the Chinese Longitudinal Healthy Longevity Survey

**DOI:** 10.1101/2020.09.25.20201970

**Authors:** Xurui Jin, Hao Zhang, Kehan Zhang, Yu Leng, Yali Zhao, Yi Zeng, Junfeng (Jim) Zhang, Yao Yao

**Affiliations:** Global Health Research Center, Duke Kunshan University, Kunshan, Jiangsu, China; Department of healthcare policy and research, Weill Cornell Medicine, New York, NY, USA; Central Laboratory, Hainan Hospital of Chinese People’s Liberation Army General Hospital, Sanya, China; Center for Healthy Aging and Development Studies, Raissun Institute for Advanced Studies, National School of Development, Peking University, Beijing, China; Center for the Study of Aging and Human Development and Geriatrics Division, Medical School of Duke University, Durham, NC, United States; Nicholas School of the Environment and Duke Global Health Institute, Duke University, Durham, NC, United States; Aladdin Healthcare Technologies Ltd, 24-26 Baltic Street West, London EC1Y OUR, UK

**Author notes:** Corresponding author:Junfeng (Jim) Zhang, Nicholas School of the Environment and Duke Global Health Institute, Duke University, Durham, NC, United States., Email address, Yi Zeng, Center for Healthy Aging and Development Studies, Raissun Institute for Advanced Studies, National School of Development, Peking University, Beijing, China., Email address, Yao Yao, Center for Healthy Aging and Development Studies, National School of Development, National School of Development, Peking University, Beijing, China. Those authors contributed equally to this work (Xurui Jin, Hao Zhang).

**Keywords:** Depressive symptoms, Natural contact, Ventilation, Window-opening, Older adults

## Abstract

**BACKGROUND:** Recent evidence suggests that window-view and window-ventilation may contribute to mental wellbeing. Compared to their younger counterparts, older adults spend more time at home and have less contact with natural environments due to social isolation and physical decline. However, the association of residential window-opening with depressive symptoms is understudied among older populations.

**METHODS:** We used data from a community-based cohort study conducted in 23 provinces of China including 13,125 adults age 65 years or older (mean age: 83.8 years [SD: 11.1]). We created the window opening index (WOI) as a proxy of window-view and window-ventilation, leveraging relevant data on self-reported frequencies of manual window opening at home. Depressive symptoms were assessed by the 10-item version of the Centre for Epidemiological Studies Depression scale no fewer than 10. We used multivariate logistic regression model to examine the association between window opening and depressive symptoms.

**RESULTS:** In the multivariate logistic regression model controlling for a set of well-designed mood-influencing environmental and individual-level covariates, a protective effect of window opening on depressive symptoms was observed, with 16% lower odds per interquartile increment in WOI (odds ratio: 0.84, 95%-CI: 0.81-0.87). Subgroup analyses indicated that the beneficial effects were more pronounced among those with higher socioeconomic status (higher levels of education, urban residents) and better surrounding environments (lower exposure of ambient fine particular matter, proximity to park, or higher levels of residential greenness).

**DISCUSSION:** The results point to the benefits of well-designed household window-opening environments on the mental health of older populations and suggest a synergistic effect of window-opening and favorable surrounding natural environment.

## Introduction

Mental disorder weighs a substantial proportion of the burden of maladies around the world. The global burden of mental illness accounts for 32.4% of years lived with disability (YLDs) and 13.0% of disability-adjusted life-years (DALYs) (Vigo et al. 2016). Older adults are more susceptible to mental health problems due to social isolation, loneliness, and physical frailty (Blazer 2020). Among this population, mental disorder such as depression is associated with an increased risk of all-cause and cardiovascular mortality, chronic conditions, dementia as well as suicidal ideation, attempts and death (Fiske et al. 2009; Ribeiro et al. 2018).

A variety of factors have been identified to affect mental health, including but not limited to social, economic, psychological, physiological, behavioral, environmental, genetic, and epigenetic influences (Meyer-Lindenberg 2014). Recent studies have prompted attention to the potential anti-depressant effect of nature contact (Bratman et al. 2019; Kruize et al. 2020; Sarkar et al. 2018). Definition of nature contact was derived from both physical experiencing (i.e. wilderness adventures, outings in natural settings, gardening, visiting a public garden, land care activity) and nature viewing (i.e. window view of nature, viewing images of natural landscapes) (Frumkin et al. 2017). Window opening is connected to two forms of nature experiences: the viewing of nature and the ventilation of indoor air. However, it is still unknown if and to what extent viewing nature through window can contribute to the mental wellbeing among older population.

Previous studies have brought to light several benefits of regular window opening. For example, nature viewing from window can boost recovery from a surgery (Ulrich 1984) and elicit improvements in the recovery process following a stressor (Brown et al. 2013). A recent study reviewed thirty-seven articles and concluded that viewing nature has physiological benefits (Jo et al. 2019). Another benefit of window opening is that indoor air exchanging, also defined as natural ventilation (Escombe et al. 2007), could directly or indirectly affect health through following pathways: lowering the concentrations of indoor air pollutants (Tong et al. 2016), managing indoor temperature and moisture levels (Francisco et al. 2017; Ye et al. 2017), as well as reducing the risk of airborne contagion such as tuberculosis (Escombe et al. 2007) and coronavirus disease 2019 (COVID-19) (Chen and Zhao 2020). In addition, window opening has protective effects on cognitive functions (Ko et al. 2020) and can also decrease the risk of sick building syndrome (i.e. a medical condition displaying respiratory symptoms and mental illnesses such as headache, fatigue, difficulty in concentration, and personality changes) (Fisk et al. 2009; Fisk 2018). However, the majority of the studies focused on the natural impact of window opening on mental wellbeing among the young and middle-aged population, while little is known about the impact on older adults. Since older adults are more likely to spend time at home than their younger counterparts due to retirements, social isolation, loneliness, and physical frailty (Blazer 2020), it is important and intriguing to understand whether household window opening, an indicator of window-view and window-ventilation, can improve the mental health for older population.

The present study aimed at exploring the association of household window opening with depressive symptoms, through the analysis of a nationwide dataset of community-dwelling older adults in China. Considering that the health effects of window opening can be realized not only through nature viewing but also ventilation, said effects might also be influenced by relevant covariates such as urban residency, socioeconomic status, and surrounding environmental factors (Bratman et al. 2019; Frumkin et al. 2017; Sarkar et al. 2018). The present study did further analyses stratified by age, sex, urban residency, ambient air pollution exposure, residential greenness exposure, proximity to the nearest park, and polluted fuel use.

## Methods

### Study population

The present study used the data from the 2018 wave of CLHLS, a longitudinal study since 1998 with follow-up surveys every 2 to 3 years. The CLHLS randomly selected 806 cities and counties in 23 provinces of China using multi-stage stratified sampling. The study areas covered 85% Chinese population. More details on sampling design and data quality can be found elsewhere (Zeng et al. 2017a; Zeng et al. 2017b).

The 2018 wave of CLHLS had a total of 15,200 participants aged 65 and older. After excluding 2075 participants due to missing data on the 10-item Center for Epidemiologic Studies Depression Scale (CES-D-10), the final analytical sample included 13,125 participants (5,080 aged 65-79 years old and 8,075 aged ≥80 years old). The participant flow chart can be seen in Appendix Figure S1. The design of CLHLS was approved by the Campus Institutional Review Board of Duke University (Pro00062871) and the Biomedical Ethics Committee of Peking University (IRB00001052-13074). All participants or their legal representatives signed written consent forms during the surveys.

### Assessment of household window opening

The data on self-reported seasonal frequencies of window opening was collected by trained research staffs through personal interviews. We created the window opening index (WOI) as a proxy of window-view and window- ventilation, leveraging the relevant data on self-reported frequencies of manual window opening at home. A 3- linkert scale question “In the last 12 months, how often did you open the window manually for nature view and ventilation” was used to measure the manual window-opening frequency in each season: ‘0 to 2 time per week’, ‘3 to 5 times per week’, ‘>5 times per week’. Those three terms received a score of 0, 1, or 2, respectively, with a higher item score indicated higher frequency of viewing nature. The annual WOI was then summed with those scores from four seasons (spring, summer, fall, and winter) and ranged from zero to eight. We categorized the annual WOI into low (WOI: 1, 2), low-medium (WOI: 3, 4), high-medium (WOI: 5-7) and high groups (WOI: 8) by quartiles for our further analysis.

### Assessment of depressive symptoms

We used the CES-D-10 to measure depressive symptoms. The answers were indicated using a four-level scale: “rarely” (0-1 day), “some days” (1-2 days), “occasionally” (3-4 days), and “most of the time” (5-7 days). For the two positive questions— “I was happy” and “I felt hopeful about the future”, answers were reversely coded before summation. We then coded all answers from “rarely”, “some days”, “occasionally” to “most of the time” as 0 to 3, respectively. The CES-D-10 score in this study ranged from 0 to 30, with higher scores indicating greater severity of depressive symptoms. A person is considered to have depressive symptoms if they scored no fewer than 10 in the CES-D-10. This threshold of 10 has been widely applied in previous studies and well validated in the measurement of depressive symptoms in Chinese older populations (Cheng and Chan 2005, 2008).

### Mood-influencing environmental covariates

We considered an array of environmental covariates, following the existing literature on potential risk factors for mental health, including ambient fine particulate exposure, residential greenness exposure, proximity to the nearest park, polluted fuel use, and use of kitchen range hood (Banay et al. 2019; Gu et al. 2019; Liu et al. 2020; Mukherjee et al. 2017; Sarkar et al. 2018).

Based on the participants’ residential addresses, the ground-level concentrations of PM_2.5_ were estimated from the Atmospheric Composition Analysis Group (van Donkelaar et al. 2015). The details on the estimation of PM_2.5_ concentration can be referred to previous published papers (Li et al. 2018; Xue et al. 2019a). Ground-level PM_2.5_ concentrations were estimated by combining aerosol optical depth retrievals from multiple satellite products (MISR, MODIS Dark Target, MODIS and SeaWiFS Deep Blue, and MODIS MAIAC) and then calibrating to ground-based observations of PM_2.5_ using geographically weighted regression (Li et al. 2018; Xue et al. 2019a). Annual PM_2.5_ estimates were calculated in 2018, at 1 km^2^ spatial resolution, which was the longest and the highest resolution exposure dataset available (Li et al. 2018). In addition, our estimates were highly consistent with out-of-sample cross-validated concentrations from monitors (R^2^=0.81) and another exposure dataset in China (R^2^=0.81) (Ji et al. 2020; Li et al. 2018).

As for the residential greenness, we used remote sensing to calculate the normalized difference vegetation index (NDVI) around participants’ addresses. We measured NDVI values from the Moderate-Resolution Imaging Spectro-Radiometer (MODIS), with a varying spatial resolution up to 500 m, in the National Aeronautics and Space Administration’s Terra Satellite (Thiering et al. 2016). In line with suggestions of previous studies (Ji et al. 2020; Sarkar et al. 2018), we use NDVI in the 500 m radius to represent the surrounding greenness of the respective participants in our study. We calculated the annual greenness exposure by averaging surrounding NDVI values throughout the one year before the survey. For example, if a participant was surveyed on July 1, 2018, we then took the average value of the 12 months NDVI (from July 1, 2017 to July 1, 2018). The details of the residential greenness ascertainment were described in previous studies (Ji et al. 2020; Xue et al. 2019b).

We calculated the measures of distance to the nearest park using ArcGIS 10.3. A park was defined as “a public park or outdoor recreation area in the community that is designed for active or passive use” (Bai et al. 2013). We used street network distance to estimate the distance from the participant’s residency to the geometric centroid of the nearest park, a method that was proved to be more appropriate in simulating walking behavior than Euclidean distance (Apparicio et al. 2008; Witten et al. 2003). Although there is no available common standard readily defining proximity to the nearest parks, a 1 mile-buffer (approximately 1600 m) was thought to be a reasonable walking distance and is similar to the ones used in several previous studies of physical activity and park facilities (Kaczynski et al. 2014). Considering that the participants in our study were older adults (mean age of 83.8 years old) and nearly one-quarter of them were physically inactive (22.7%), we used a distance of 0.5 mile (800 m) as the cutoff buffer value to reflect the availability of park facilities.

Participants were asked to report the main type of cooking fuel they used during the past 5 years. In line with previous studies, polluted fuel included coal, biomass charcoal, woods or straw, and clean fuel consisted of natural gas fuel, liquified gas or electricity (Deng et al. 2020; Liu et al. 2020).

### Individual-level covariates

In addition to the potential environmental covariates, we also included a wide range of individual-level confounding factors in our analysis, consisting of demographic information (sex, age, marital status, residency, city population, geographical region), socioeconomic status (education level, household annual income, medical insurance coverage), lifestyle factors (tobacco smoking, alcohol consumption, dietary diversity, physical activity), and health-related factors (body mass index [BMI], self-rated health, cognition, and activity of daily living [ADL]). The individual-level covariates were defined as followed: residency (rural vs. urban), education level (none vs. any [≥1 years of schooling]), marital statues (currently married and living with spouse vs others), tobacco smoking (current vs. not current), alcohol consumption (current vs. not current), physical activity (current vs. not current), body mass index, cooking fuel (clean [electricity, gas, solar energy] vs. polluted [charcoal, firewood /straw]), family income (“<30,000 RMB/yr” vs. “≥30,000 RMB/yr), city population (>8 million” vs. “≤8 million), PM_2.5_ level (<55 ug/m^2^ vs. ≥55 ug/m^2^), use of kitchen range hood (never vs. sometimes or often), residential proximity to the nearest park (>800 meters vs. ≤800 meters), geographical region (Northern China [Beijing, Tianjin, Hebei, Shanxi, Shaanxi, Shandong, Liaoning, Jilin, and Heilongjiang provinces], Eastern China [Shanghai, Jiangsu, Zhejiang, and Fujian provinces], Central China [Henan, Hubei, Jiangxi, Anhui, and Hunan provinces], Southwestern China [Guangdong, Guangxi, Chongqing, Sichuan, and Hainan provinces]), participation of two kinds of medical insurances (Urban employee/resident medical insurance, New rural cooperative medical insurance), cognitive impairment (defined by MMSE score<25; with vs. without), physical disability for activity of daily living (with vs. without).

### Statistical Analysis

Missing data for the covariates at baseline accounts for less than 1.0% and multiple imputation methods were adopted to impute those missing values. Logistics regression models were performed to examine the association of WOI with depressive symptoms, adjusting for mood-influencing environmental (ambient fine particulate exposure, residential greenness exposure, proximity to nearest park, polluted fuel use, and use of kitchen range hood) and individual-level (sex, age, marital status, residency, city population, geographical region, education level, family annual income, medical insurance coverage, tobacco smoking, alcohol consumption, dietary diversity, physical activity, BMI, self-rated health, cognition, and ADL) covariates. Multivariate logistic regression with penalized splines evaluated the non-linear associations of WOI with depressive symptoms.

Furthermore, we also conducted additional subgroup and interaction analyses, where WOI was dichotomized into two groups (highest and second highest vs. lowest and second lowest). We examined whether the association between window opening and depressive symptoms differed by eleven predefined factors: sex, age (<80 years old versus ≥80 years old), education level, urban/rural residency, tobacco smoking, use of kitchen range hood, indoor cooking ventilation, PM_2.5_ level, city population, residential proximity to the nearest park, and geographical region. The significance of multiplicative interactions between dichotomized WOI and other variables was tested by cross-product terms in the models.

Additionally, we performed several steps of sensitivity analyses to assess the robustness of the study results. First, we repeated our analyses using varied cut-off values of the CES-D-10 such as 8 and 12. Second, we excluded participants with severe cognitive impairment with scores of MMSE < 18 (Tombaugh and McIntyre 1992), among whom substantial recall bias might have occurred. Third, we excluded participants who changed their residential addresses within five years and focused on non-movers (Tzivian et al. 2016). Fourth, we tested our results using the data before multiple imputation and further adjusted sampling weight. Lastly, our model was additionally adjusted for seven self-reported chronic conditions (hypertension, diabetes, heart disease, stroke, chronic pulmonary disease, arthritis, and cancer). All analyses were performed using STATA version 16.0 (Stata Corp, College Station, TX, USA).

## Results

### Demographic characteristics

A total of 13,125 participants were included in the present study with a mean age of 83.8 (SD=11.1) years; 7,105 (54.1%) were women and 7,300 (55.6%) were urban residents (Table 1). Among them, 56.5% of the study participants had a CES-D-10 score ≥10, indicating depressive symptoms. Overall, 2,941 (22.4%), 2,534 (19.3%), 2,868 (21.9%) and 4,782 (36.4%) participants had low, low-medium, high-medium, and high frequencies of WOI, respectively. Participants having higher frequencies of household window opening were more likely to be younger, had higher education level and family income, lived in urban areas, lived in areas with lower PM_2.5_, were proximate to parks, were smokers, had higher physical activity levels, were adopting range hood while cooking, and were free of cognitive impairments.

**Table 1.**
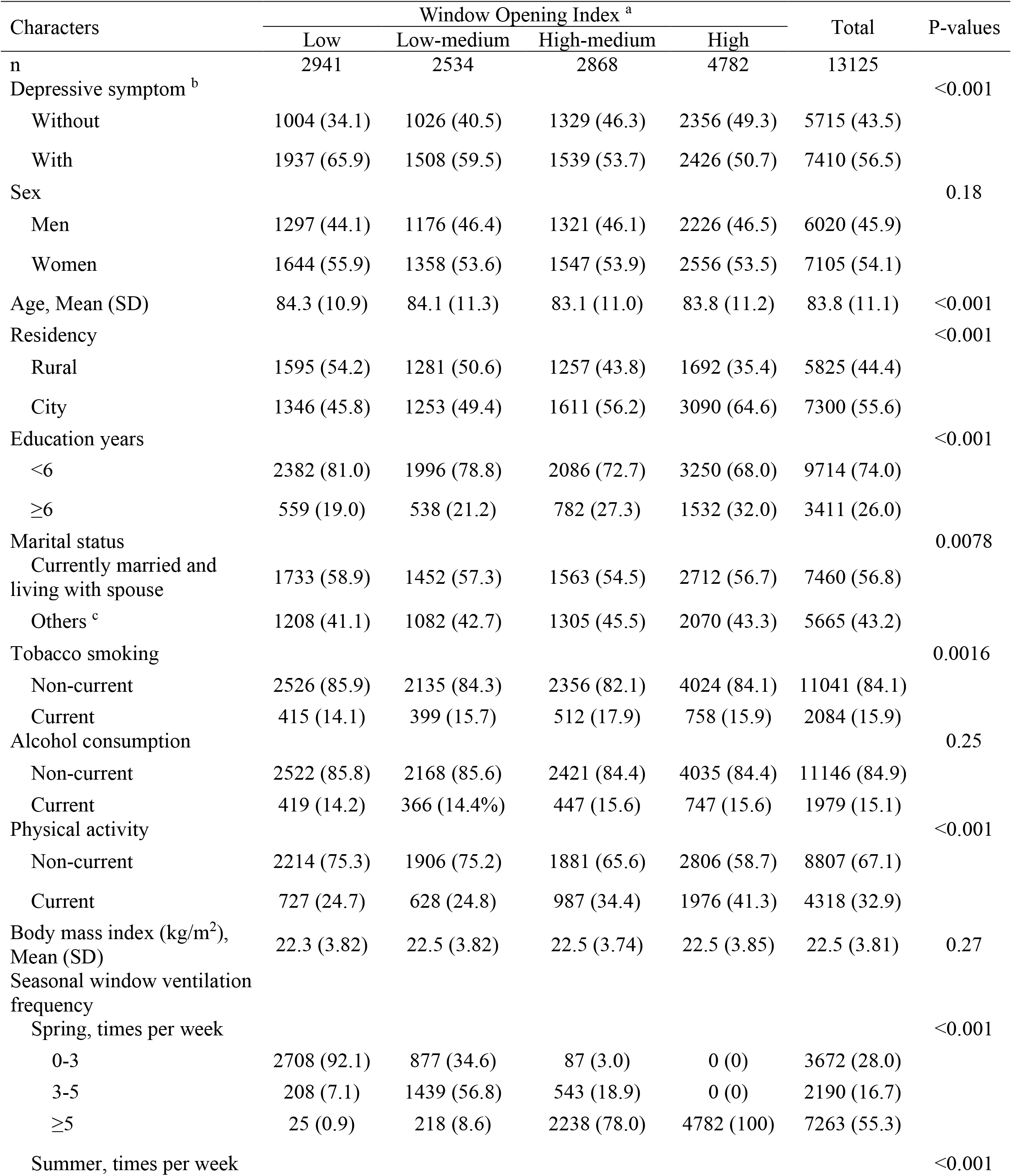
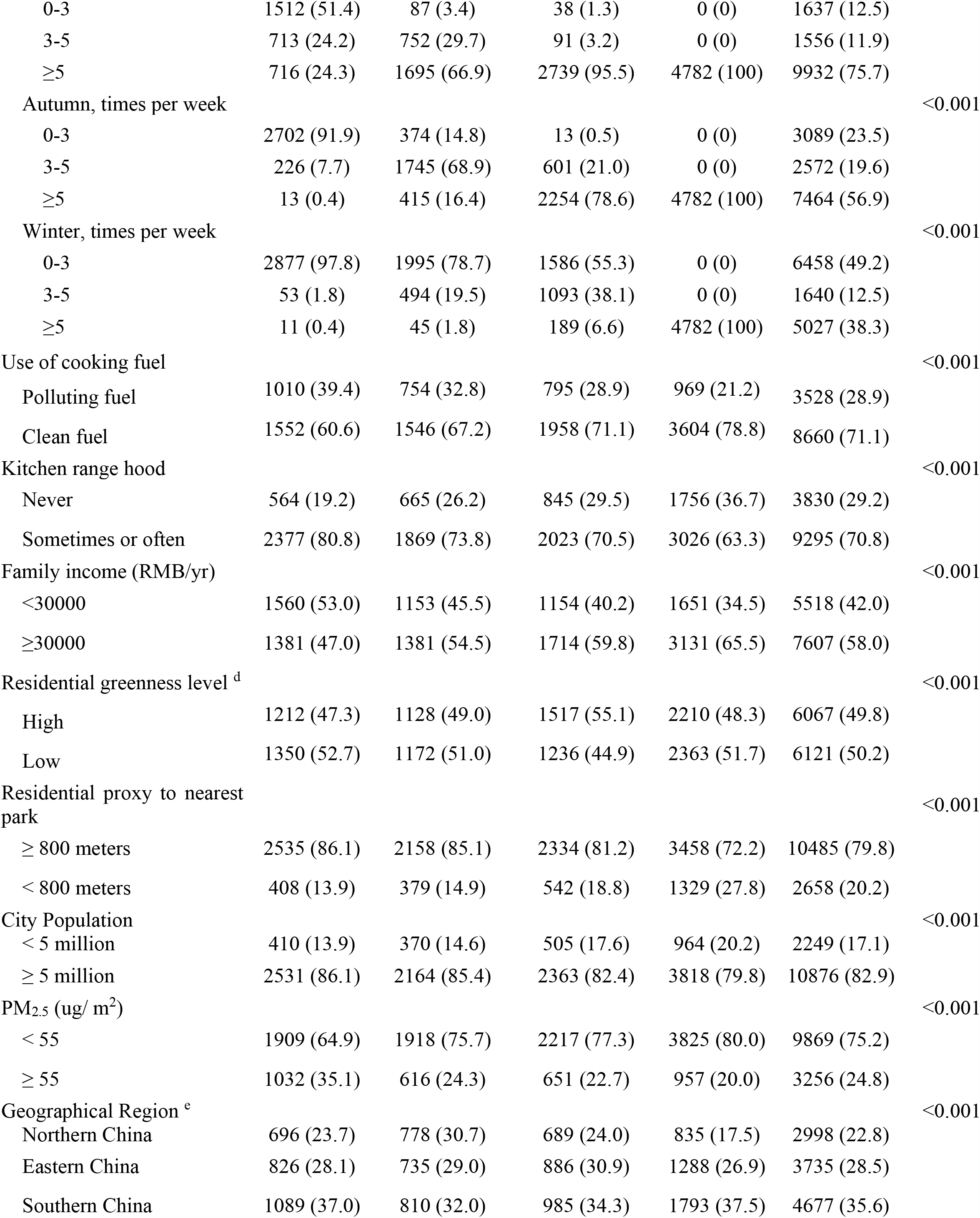
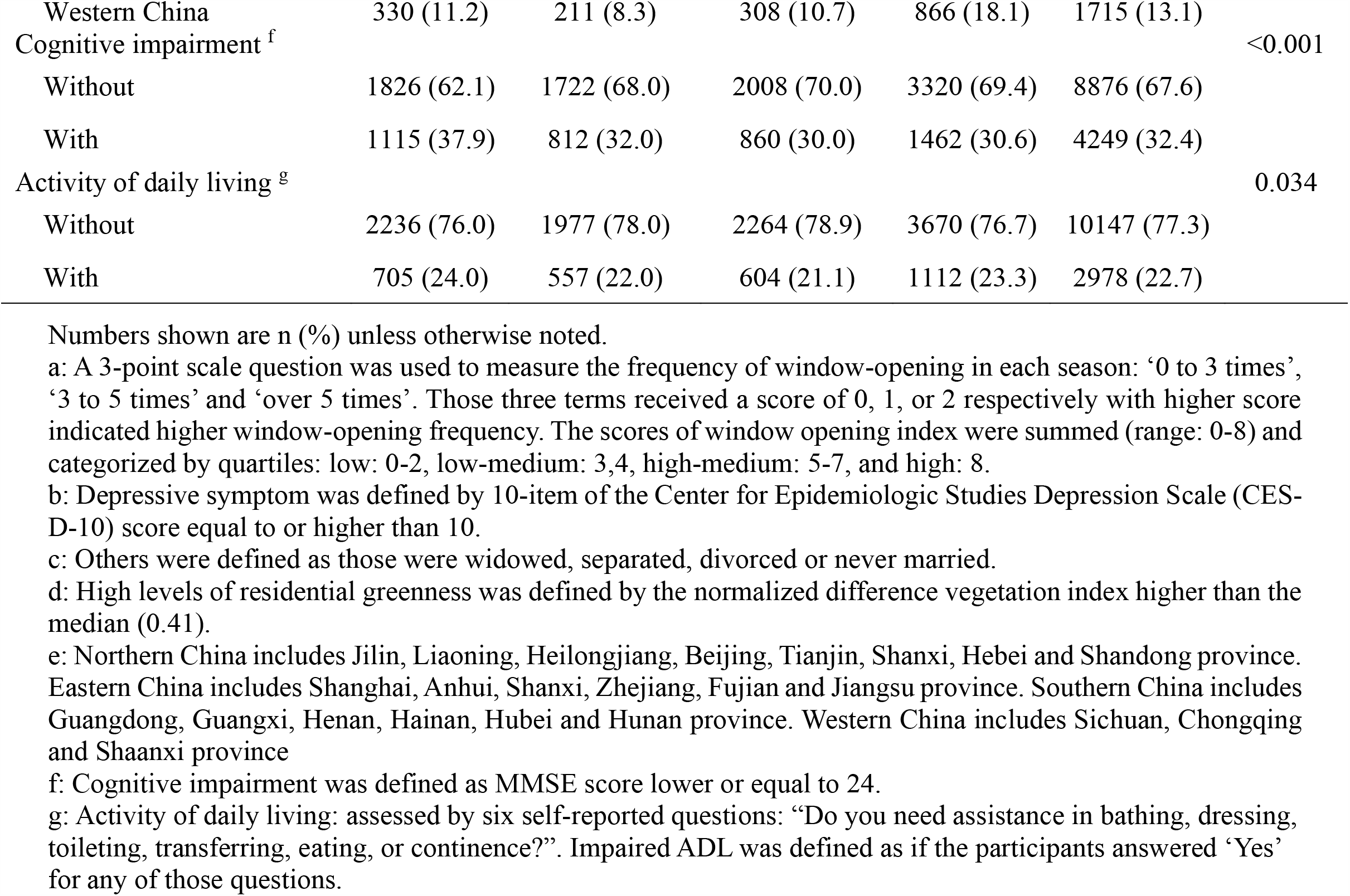
Baseline Characteristics of the 13,125 Study Participants According to the Quartiles of Window Opening Index

### Association of annual and seasonal window opening with depressive symptoms

Regarding the association of annual WOI with depressive symptoms, the fully-adjusted odds ratio (OR) of depressive symptom for persons with high-medium, low-medium and low frequencies of household window opening were 1.11 (95% CI: 1.00, 1.22), 1.40 (95% CI: 1.26, 1.56), and 1.68 (95% CI: 1.51, 1.86), respectively, compared with high frequency (Figure 1). We found a 16% lower odds of depressive symptoms per interquartile increment in WOI (odds ratio 0.84, 95% CI 0.81-0.87; *P*<0.001). The frequency of window-opening demonstrated a linear association with depressive symptoms (*P* for a linear trend<0.001), revealed in the fully-adjusted penalized spline regression (Figure 2).

**Figure 1.**
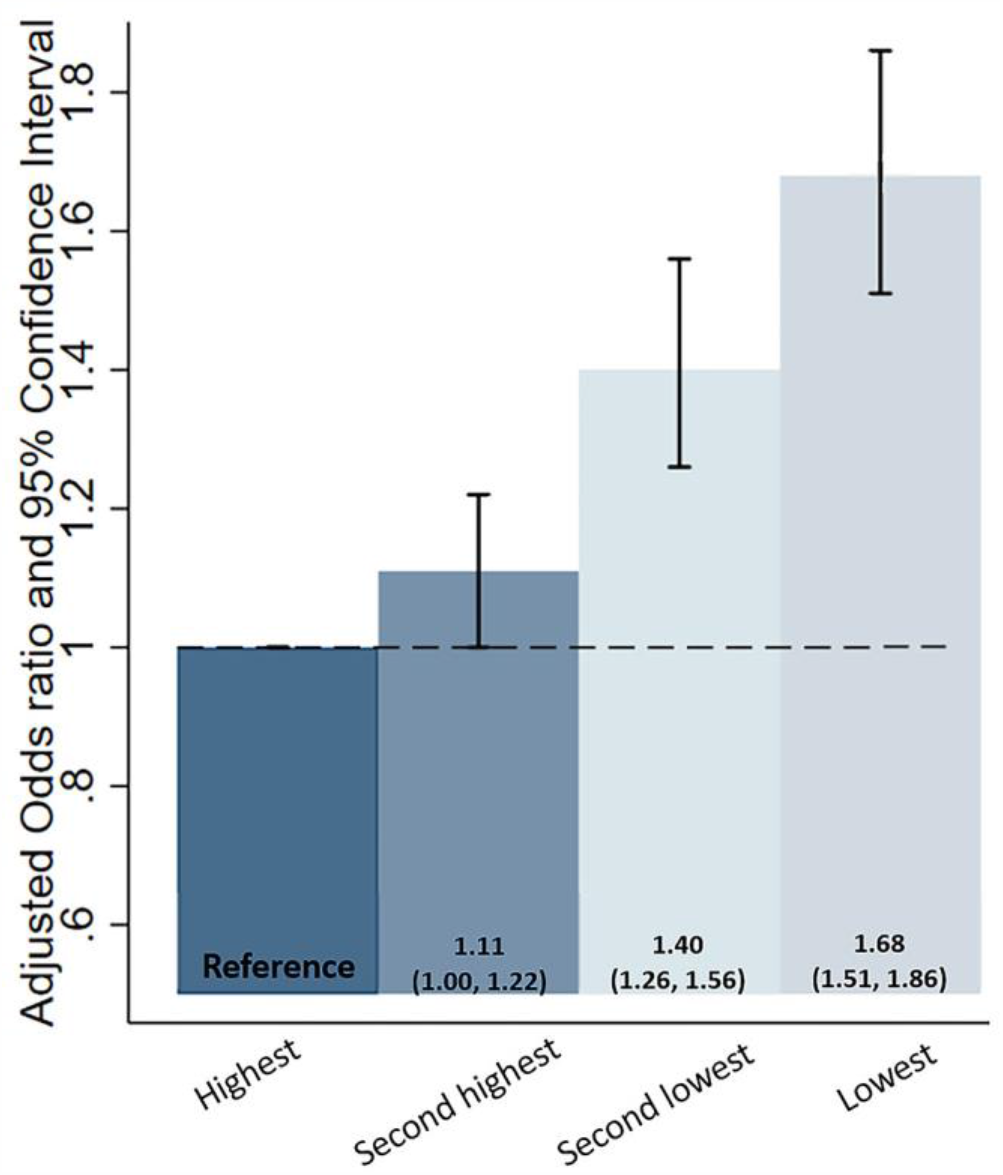
Association between Quartiles of Window Opening Frequency and Depressive Symptom. A 3-point scale question was used to measure the frequency of window-opening in each season: ‘0 to 3 times’, ‘3 to 5 times’ and ‘over 5 times’. Those three terms received a score of 0, 1, or 2 respectively with higher score indicated higher ventilation frequency. The scores of window opening index were summed (range: 0-8) and categorized by quartiles: lowest: 0-2, second lowest: 3,4, second highest: 5-7, highest: 8. Adjustments: environmental factors (ambient fine particulate exposure, residential greenness exposure, proximity to nearest park, and polluted fuel use) and individual confounders (sex, age, marital status, residency, city population, geographical region, education level, family annual income, medical insurance coverage, tobacco smoking, alcohol consumption, dietary diversity, physical activity, BMI, self-rated health, cognitive function, ADL).

**Figure 2.**
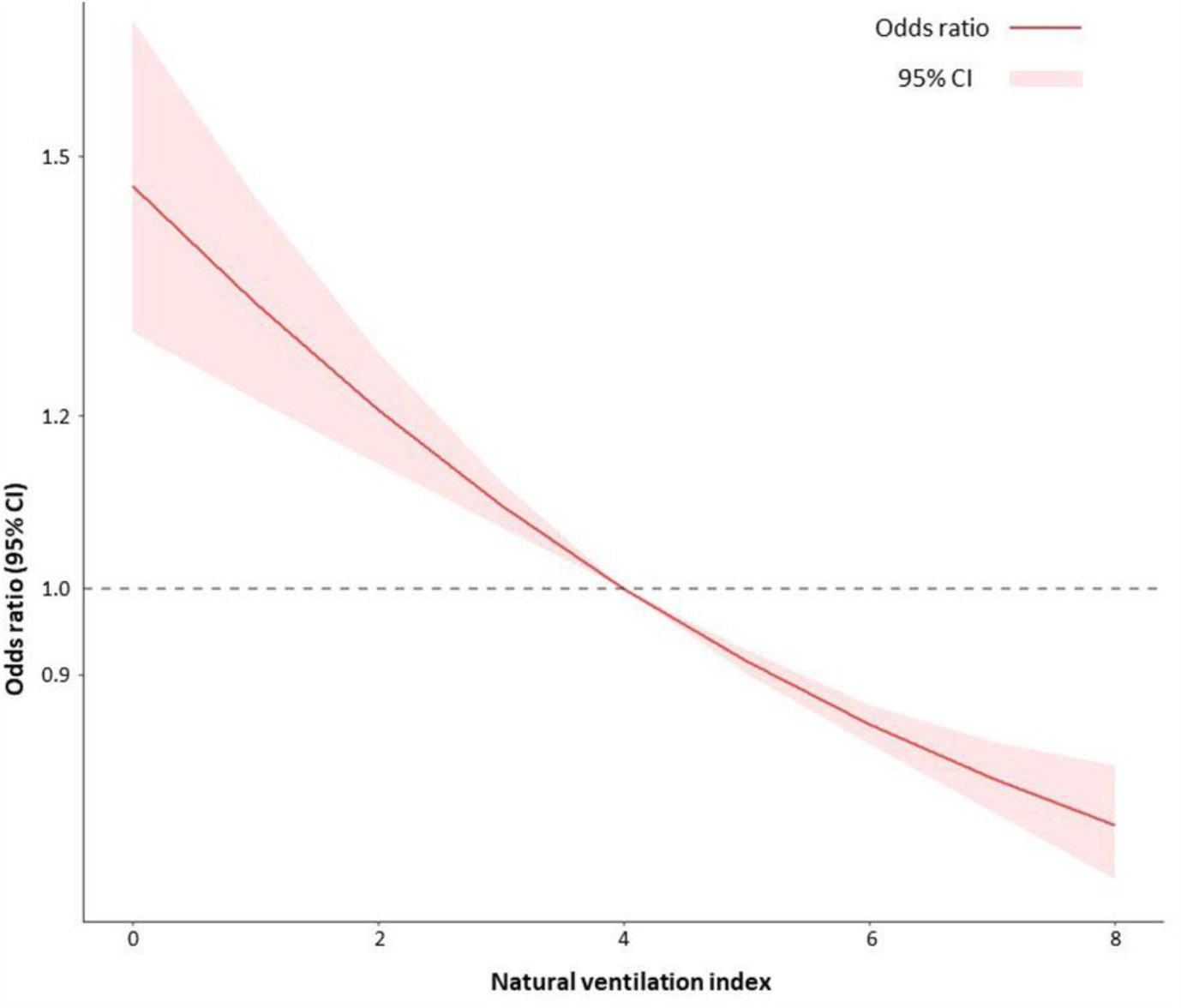
Association of Window opening Frequency and Depressive Symptom by Penalized Spline. A 3-point scale question was used to measure the frequency of window-opening in each season: ‘0 to 3 times’, ‘3 to 5 times’ and ‘over 5 times’. Those three terms received a score of 0, 1, or 2 respectively with higher score indicated higher window-opening frequency. The scores of window opening index were summed (range: 0-8). Adjustments: environmental factors (ambient fine particulate exposure, residential greenness exposure, proximity to nearest park, and polluted fuel use) and individual confounders (sex, age, marital status, residency, city population, geographical region, education level, family annual income, medical insurance coverage, tobacco smoking, alcohol consumption, dietary diversity, physical activity, BMI, self-rated health, cognitive function, ADL).

After reanalyzing specific association of WOI with depressive symptoms in each season, we found that, in spring and autumn, the prevalence of depressive symptoms was significantly lower among individuals with higher window-opening frequency (Figure S2). The odds of having depressive symptoms were 47% and 18% higher in spring and autumn, respectively (*P*s<0.05), among those with fewer than 3 times window-opening per week compared to those with more than 5 times window-opening per week. In contrast, we observed an opposite result of the association between window-opening and depression in winter, during which less frequent window-opening was associated with lower odds of depressive symptoms (<3 times/week vs. >5 time/week, OR=0.84, 95% CI: 0.75-0.94; *P*<0.05) (Figure S2).

### Effect modification and Sensitivity analysis

We conducted subgroup analyses by eleven predefined factors. Significantly larger beneficial effects of window opening were observed among those who were more educated, who were rural residents, who were tobacco smokers, who did not adopt kitchen ventilation while cooking, who lived in areas with lower level of ambient PM_2.5_, who resided more proximately to parks, and who lived in a city with higher population density (Table 2).

**Table 2.** Subgroup Analysis of Association between Window Opening Frequency and Depressive Symptom.

| Characteristic | Participants | Odd Ratio<br>(95% CI) | *P* for interaction |
| --- | --- | --- | --- |
| **Sex** |  |  | 0.15 |
| Female |  |  |  |
| High window opening | 4103 | Ref. |  |
| Low window opening | 3002 | 1.55 (1.37, 1.75) |  |
| Male |  |  |  |
| High window opening | 3547 | Ref. |  |
| Low window opening | 2473 | 1.41 (1.27, 1.58) |  |
| **Age** |  |  | 0.31 |
| ≥ 80 |  |  |  |
| High window opening | 4595 | Ref. |  |
| Low window opening | 3450 | 1.44 (1.30, 1.60) |  |
| < 80 |  |  |  |
| High window opening | 3055 | Ref. |  |
| Low window opening | 2025 | 1.51 (1.33, 1.73) |  |
| **Education years** |  |  | <0.001 |
| 0-6 years |  |  |  |
| High window opening | 5336 | Ref. |  |
| Low window opening | 4378 | 1.32 (1.20, 1.45) |  |
| >6 years |  |  |  |
| High window opening | 2314 | Ref. |  |
| Low window opening | 1097 | 1.88 (1.60, 2.21) |  |
| **Residency** |  |  | <0.001 |
| Urban |  |  |  |
| High window opening | 4701 | Ref. |  |
| Low window opening | 2599 | 1.83 (1.63, 2.04) |  |
| Rural |  |  |  |
| High window opening | 2949 | Ref. |  |
| Low window opening | 2876 | 1.14 (1.01, 1.29) |  |
| **Smoking** |  |  | 0.025 |
| Not current |  |  |  |
| High window opening | 6380 | Ref. |  |
| Low window opening | 4661 | 1.42 (1.30, 1.56) |  |
| Current |  |  |  |
| High window opening | 1270 | Ref. |  |
| Low window opening | 814 | 1.79 (1.45, 2.20) |  |
| **Kitchen range hood use** |  |  | 0.007 |
| Never |  |  |  |
| High window opening | 823 | Ref. |  |
| Low window opening | 363 | 2.05 (1.53, 2.75) |  |
| Sometimes or often |  |  |  |
| High window opening | 6827 | Ref. |  |
| Low window opening | 5112 | 1.42 (1.31, 1.55) |  |
| **PM<sub>2.5</sub> level** |  |  | 0.012 |
| < 55 |  |  |  |
| High window opening | 6081 | Ref. |  |
| Low window opening | 3672 | 1.72 (1.47, 2.01) |  |
| ≥ 55 |  |  |  |
| High window opening | 1569 | Ref. |  |
| Low window opening | 1803 | 1.38 (1.25, 1.51) |  |
| **City population** |  |  | 0.012 |
| ≥ 5 million |  |  |  |
| High window opening | 5292 | Ref. |  |
| Low window opening | 3579 | 1.28 (1.12, 1.47) |  |
| < 5 million |  |  |  |
| High window opening | 2358 | Ref. |  |
| Low window opening | 1896 | 1.62 (1.47, 1.80) |  |
| **Residential proxy to nearest park** |  |  |  |
| ≥ 800 meters |  |  |  |
| High window opening | 5784 | Ref. | <0.001 |
| Low window opening | 4689 | 1.30 (1.20, 1.43) |  |
| < 800 meters |  |  |  |
| High window opening | 1866 | Ref. |  |
| Low window opening | 786 | 2.20 (1.80, 2.69) |  |
| **Residential greenness** |  |  | 0.052 |
| High |  |  |  |
| High window opening | 3727 | Ref. |  |
| Low window opening | 2340 | 1.63 (1.45, 1.83) |  |
| Low |  |  |  |
| High window opening | 3599 | Ref. |  |
| Low window opening | 2522 | 1.34 (1.20, 1.51) |  |
| **Geographical region** |  |  | 0.033/ 0.001/ 0.007 |
| Northern China |  |  |  |
| High window opening | 1526 | Ref. |  |
| Low window opening | 1473 | 1.73 (1.46, 2.07) |  |
| Eastern China |  |  |  |
| High window opening | 2170 | Ref. |  |
| Low window opening | 1559 | 1.80 (1.54, 2.09) |  |
| Southern China |  |  |  |
| High window opening | 2781 | Ref. |  |
| Low window opening | 1903 | 1.16 (1.00, 1.34) |  |
| Western China |  |  |  |
| High window opening | 1173 | Ref. |  |
| Low window opening | 540 | 1.38 (1.10, 1.75) |  |

Adjustments: age at baseline, sex, residency, education level, marital status, tobacco smoking, alcohol consumption, physical activity, body mass index, indoor cooking ventilation, cooking fuel, family income, city population, PM<sub>2.5</sub> level, use of oil remover, proxy to the nearest park, geographical region, participation of two kinds of medical insurance (Urban employee/resident medical insurance, New rural cooperative medical insurance), cognitive impairment, activity of daily living.
High window opening frequency was defined by the window opening score lower than 5 and low window opening frequency was defined by the window opening score equal to or higher than 5.

In the sensitivity analysis using varied cut-off values of the CES-D-10 such as 8 and 12, we repeated the analysis for the fully-adjusted model. The dose-effect relationship of household window opening with prevalence of depressive symptoms mildly attenuated in analyses using both cutoffs (Table S1). After excluding participants with severe cognitive impairments (n=893), the effect estimates or the significance levels of the observed associations were not altered (Table S2). Moreover, after restricting the study population to non-movers and excluding those with changed addresses in the last five years (n=482), we found out that the results were identical to those we presented in the main text (Table S3). We also tested our results using the original data before multiple imputation (n=10,355) and further adjusting sampling weight (Table S4). Finally, we replaced the self-rated health with seven self-reported comorbidities (hypertension, diabetes, heart disease, stroke, chronic pulmonary disease, arthritis, and cancer) (Table S5). Results from the sensitivity analyses were all reasonably consistent with those from our main analysis.

## Discussion

In this cross-sectional study, we were the first to explore the association of household window opening with depressive symptoms, and its interaction with environmental factors in a nationwide sample of Chinese older adults. We found that a higher frequency of manual window-opening was associated with lower odds of depressive symptoms among Chinese older adults. Additionally, we found that this association was more pronounced among those participants who were more educated, who lived in urban regions, who were current smokers, who lived with poorer household air quality, who lived with less ambient fine particular matter exposure, who resided more proximately to parks, or enjoyed higher residential greenness (*P*s for interactions<0.05).

These findings confirmed our hypothesis that opening window, which accompanied with nature viewing and ventilation, may have a protective effect on mental health, with effect sizes being 16% lower odds of depressive symptoms per interquartile increment in WOI after controlling for a variety of covariates. For those older adults with a habit of higher frequency of window-opening, they were more likely to experience stronger and more lasting sensory stimuli from the external environment and the protective effect of visual connection to nature on mental health may be one possible explanation for our findings. Nature viewing has been demonstrated to have a positive impact on attention restoration, stress reduction, and overall health and well-being. A previous randomized study has reported that nature viewing (composed of trees, grass, fields), rather than urban sceneries (composed of man-made surroundings, lacking in natural characteristics), could positively affect recovery process following a stressor (Brown et al. 2013). One small-scale experimental study found a significant positive effect of the visual landscape (Lee 2017). Another experimental study, by implementing a novel neuroscience approach using functional magnetic resonance imaging (fMRI), supported the psychologically restorative effect of viewing the natural environment (Tang et al. 2017). A recent study also highlighted the beneficial effect of window ventilation for emotion and mood, and it showed that more positive emotions (e. g., happy, satisfied) and fewer negative emotions (e.g., sad, drowsy) for the participants in the window versus the windowless condition. It also suggested that providing a window with a view might boost the comfort, emotion, memory and concentration for the occupants (Ko et al. 2020). Additionally, the benefits of higher levels of natural viewing through window-opening might be attributed to the mechanisms of biology, physiology, and lifestyle. From a biological perspective, the biophilia hypothesis suggested that humans are intrinsically affiliated with other forms of life including natural environment (Kellert and Wilson 1993), and viewing such environments are inherently associated with lower level of stress and tension than urban environments (Bratman et al. 2019; Ko et al. 2020). In addition, viewing natural environments through windows can reduce the stress-related biomarkers such as cortisol and oxygenated hemoglobin (O_2_Hb) (Lee 2017) and is associated with lower frequency of frontal cortex activity (Norwood et al. 2019). From a physiological standpoint, a recent systematic review found that viewing natural scenery led to more relaxed body responses by measuring several physiological indicators (functional magnetic resonance imaging [fMRI], near-infrared spectroscopy [NIRS] and electroencephalography [EEG]), and autonomic nervous activities (heart rate variability [HRV], heart rate, pulse rate and blood pressure) (Jo et al. 2019). Nature-viewing can induce higher level of HRV, reduce blood pressure and improve mood and self-esteem, by altering autonomic activity, specifically parasympathetic activity (Gladwell et al. 2012). It should be stressed that, in our study, the association between window-opening and depressive symptoms was more pronounced among those who resided closer to the nearest park or were surrounded with higher residential greenness. Our findings were in line with prior studies which demonstrated that the viewing of nature-themed urban park sceneries relieved stress and restored attentional levels (Wang et al. 2016). In addition, extant findings also partially supported that the observed benefits of window opening might be explained by the higher levels of nature viewing.

Another plausible explanation of our findings could be the natural ventilation from window opening (Fisk 2018; Ko et al. 2020; Najafi et al. 2019). Opening window can increase the indoor infiltration rates and reduce prevalence rates of sick building syndrome (SBS) symptoms (Fisk et al. 2009; Fisk 2018). Adequate indoor ventilation helps prevent the accumulation of harmful environmental threats such as pollutants from building materials, fuel burning, and radon gas emissions, as well as molds, a risk factor for allergies and asthma, and vectors which may carry transmittable viruses (WHO 2020). Natural ventilation is regarded as a low-energy and climate friendly housing measure for increasing air exchanges and air quality, thereby reducing risks of noncommunicable (i.e., cardiovascular disease, stroke, injuries, asthma and other respiratory diseases) and communicable (i.e., airborne and waterborne infectious diseases, and vector-borne diseases) diseases. Specifically, it was estimated that improved natural ventilation could reduce lung-related illnesses by up to 20% (WHO 2020). Studies also suggested that natural ventilation is mostly achieved by walking to the window and manually opened it; during this process, one would more or less breathe in the refreshing air from the window, which could further help improve the mental health based on previous findings (Brown et al. 2013; Jo et al. 2019; Norwood et al. 2019). In our study, the participants less frequent in window opening who also lived with poorer household air quality had higher prevalence of depressive symptoms compared to those with a habit of window opening. It is plausible that poor indoor air quality and low frequency of window-opening would facilitate the accumulation of pollutants in the living space and then jointly increased the risk of depressive symptoms.

We also found a seasonal variation on the association between household window opening and depressive symptoms. Similar associations were found in spring and autumn while the association in winter was in the opposite direction. The observed negative association in winter may be attributed to less greenness from the view and the infiltration of outdoor air pollution (Carlton et al. 2019) or cold temperature (Chan et al. 2018).

We also observed some synergistic effects of window opening and residential environments such as greenness, proximity to parks, and ambient air pollution. Previous research reported that exposure to residential greenness was associated with better mental health (Banay et al. 2019; Sarkar et al. 2018). Frequent window opening can enhance the contact with nature through viewing and ventilation, which may partially explain the synergistic effects of household window opening and more residential greenness exposure, favorable ambient air quality, and proximity to the nearest park, on mental health in our study. A similar study based on CLHLS also observed that residential greenness and PM_2.5_ exposure interacted non-linearly with mortality and postulated that the effect of reducing air pollution might be one mechanism through which greenness can protect health (Ji et al. 2020). Our study contributed to the current body of research by showing that residential environments and window opening, by means of enhanced nature contact through window viewing and ventilation, may synergistically affect human health.

## Strengths and Limitations

Our study has several strengths. First, we used a nation-wide, large sample of Chinese older adults, and we implemented well-validated screening tools to evaluate the status of depressive symptoms. Second, our model was adjusted for a variety of mood-influencing environmental metrics suggested by the existing literature. Those environmental metrics can also help further explore the potential interactive effects between residential environments and nature contact by window-view and window-ventilation. Third, our model included a comprehensive list of individual-level covariates, consisting of demographic information, socioeconomic status, lifestyle factors, and health metrics, which made possible the identification of the vulnerable population (Sarkar et al. 2018). Finally, we performed several steps of sensitivity analyses to test the consistency and robustness of our results.

This study also has several limitations. The cross-sectional study design limited the determination of causal relationship between window opening and depressive symptoms; however, we estimated the effect size by adjusting for a number of mood-influencing environmental and individual-level covariates such as sociodemographic factors and chronic diseases, and largely excluded the residential confounding effects. In addition, the measurement of nature viewing and ventilation was self-reported, which might result in some degree of measurement errors and recall biases. We excluded participants who were regarded as suffering from mild or severe cognitive impairments and the results remained consistent. Third, the data we used did not provide us with the key information regarding clinical diagnosis for depression among the participants, limiting the clinical relevance; however, the CES-D-10 we used to measure depressive symptoms is a well-validated instrument for depressive symptoms screening among community-dwelling Chinese older populations, and was widely used in well-known nationwide surveys including National Health and Nutrition Examination Survey (NHANES), National Social Life, Health and Aging Project (NSHAP), China Health and Retirement Longitudinal Study (CHARLS) and Chinese Longitudinal Healthy Longevity Survey (CLHLS) (Carleton et al. 2013; Chen and Mui 2014; Cheng and Chan 2005; Pun et al. 2017).

## Conclusions

This study showed that the frequency of window-opening at home was significantly associated with lower odds of depressive symptoms, and suggested a synergistic effect of nature contact on mental health through an favorable residential natural environment. Our results demonstrated potential mental health benefits of habitual nature viewing and ventilation for older residents. Our study has public health implications for the increasing older populations worldwide, suggesting an importance to take well-designed window environments and window opening lifestyles into account for public health initiatives. We call for future studies to investigate a longitudinal cohort to further explore the causality between window-opening and depression, additionally adjusting for more detailed pollutants and geographic features.

## Data Availability

The data is avaliable from Peking University Open data plantform [https://opendata.pku.edu.cn/]

## Acknowledgments

This study was supported by the National Key R&D Program of China (2018YFC2000400), National Natural Sciences Foundation of China (81903392, 81941021), and China Postdoctoral Science Foundation funded project (2019M650359). The funders had no role in the design and conduct of the study; collection, management, analysis, and interpretation of the data; preparation, review, and approval of the manuscript; or the decision to submit the manuscript for publication.

